# RGC-TBRS Personal Air Pollutant Exposure versus Health Condition and Perception Pilot Study for Young Asthmatics in Hong Kong (2019/2020)

**DOI:** 10.1101/2021.06.22.21259358

**Authors:** Kenyon Chow, Yang Han, Victor OK Li, Jacqueline CK Lam, So-lun Lee, Wilfred Wong, Yu-lung Lau

## Abstract

This is a report of the RGC-TBRS funded observational pilot study which examines the effects of personal exposures to three types of air pollutants, namely, PM_1.0_, PM_2.5_, and PM_10_, on personal health condition and perception of young asthmatics (aged 12 – 15) in Hong Kong. This is the first study to investigate the relationship between PM_1.0_ and FEV_1_ and FVC of young asthmatics in Hong Kong, based on personal exposures obtained from portable sensors. Our preliminary results show that a higher level of PM_1.0_, PM_2.5_ and PM_10_ would deteriorate the health conditions of young asthmatics in HK. All correlations between particulates and lung functions are significant and negative, including PM_1.0_ exposure vs. FEV_1_ (R^2^=12%; *p*=0.023), PM_1.0_ exposure vs. FVC (R^2^=15%; *p*=0.010), PM_2.5_ exposure vs. FEV_1_ (R^2^=13%; *p*=0.019), PM_2.5_ exposure vs. FVC (R^2^=16%; *p*=0.008), PM_10_ exposure vs. FEV_1_ (R^2^=14%; *p*=0.012), and PM_10_ exposure vs. FVC (R^2^=18%; *p*=0.005). Moreover, after accounting for covariates, including age, gender, body mass index (BMI), temperature, and relative humidity, we found a significant relationship between PM_1.0_ exposure vs. FVC (Coefficient=-0.1224; *p*=0.032), PM_2.5_ exposure vs. FVC (Coefficient=-0.1177; *p*=0.021), PM_10_ exposure vs. FEV_1_ (Coefficient=-0.0703; *p*=0.019), and PM_10_ exposure vs. FVC (Coefficient=-0.1204; *p*=0.006). Further, using the pilot study data, we have performed a power analysis to estimate the sample size for our follow-up main study. Based on the primary null hypothesis that personal PM exposure would not change the FEV_1_ and FVC of young asthmatics in HK, the lowest sample size that gives 80% power at a 5% significance level is 107. Hence, the sample size (or the total number of participated asthma subjects) expected for the follow-up longitudinal clinical study should be 125 (after adjusting for the non-compliance and withdrawal of subjects). Our pilot study has demonstrated the feasibility of research into the effects of personal air pollutant exposure on health condition and health perception. Our follow-up study will address the challenges identified in the pilot study, based on the proposed follow-up actions for subject engagement, data collection, and data analysis.

## 1. Introduction

Previous air pollution and public health studies conducted internationally and in Hong Kong (HK) showed that exposures to outdoor air pollutants are correlated with the health conditions of young asthmatics. Panel studies provided the preliminary evidence for the cumulative effect of air pollutants on health conditions over time. A recent review of over 30 relevant panel studies showed that the adverse effects of air pollutant exposures on health conditions are more pronounced in asthmatic children than in healthy children [1, 2, 3, 4]. However, three major gaps have been identified from the previous studies. First, most of these panel studies are limited to the US and European contexts. Second, previous exposure assessment methods tend to use aggregate rather than personal air pollutant exposure data [5]. Further, the relationship between air pollutant exposures and health perceptions of the asthmatics is often overlooked. Until now, two important research questions are yet to be fully addressed: (1) Would a higher level of personal air pollutant exposures result in poorer health conditions among young asthmatics in HK? (2) Would a higher level of personal air pollutant exposures result in more negative health perceptions among the young asthmatics in HK?

This study aims to fill the above research gaps and investigate how personal air pollutant exposures will affect health conditions and perceptions. A pilot study was conducted during Sep 2019 – Oct 2020. 51 asthma subjects were shortlisted, and nine subjects participated in the study and finished their pilot runs. This report summarizes the results of the pilot study. The report is organized as follows. Section 2 details the data collection methodology and statistical analysis. Section 3 presents the preliminary results based on the pilot run data, compares the results with the previous literature, and calculates the sample size for our follow-up study. Section 4 lists the major challenges of the pilot study. Section 5 proposes recommendations for our follow-up study. Section 6 concludes the pilot study.

## 2. Data and Methodology

### 2.1 Data Collection during the Pilot Study

The pilot study was conducted on a rolling basis during Sep 2019 – Oct 2020. The subject recruitment was conducted by the Department of Pediatrics in Queen Mary Hospital (QMH). The pilot study targeted young asthma subjects aged 12 to 17. As of mid-Oct 2020, 51 asthma subjects were shortlisted, and nine asthma subjects (aged 12 to 15) participated in the study and finished their pilot runs. After subject enrolment, demographic information was collected through a one-off background survey (Table 1a). During the pilot study, for each subject, personal air pollutant exposure, health condition, and health perception data were continuously collected with a portable air pollution sensor, three portable/wearable devices (e-inhaler, e-spirometer, and Mi-Band), and a set of validated health-related surveys (Tables 1b-1d).

**Table 1.** Data collection frequencies and types

**(a) Demographic information**
| **Instrument** | **Sampling frequency** | **Data format** | **Data types** |
| --- | --- | --- | --- |
| Paper-based survey | Once | CSV * | Demographic information such as age, gender, weight, and height |
| **Notes:**<br>* A CSV file is a comma-separated values file that stores data in a tabular format. |  |  |  |

**(b) Personal air pollutant exposure**
| **Instrument** | **Sampling frequency** | **Data format** | **Data types** |
| --- | --- | --- | --- |
| Portable sensor * | 1 minute | CSV | Time, date, GPS longitude, GPS latitude, GPS altitude, temperature, relative humidity, background noise, three-axis accelerations, PM<sub>1.0</sub>, PM<sub>2.5</sub>, PM<sub>10</sub> |
| **Notes:**<br>* PM<sub>1.0</sub>, PM<sub>2.5</sub>, and PM<sub>10</sub> measurements are mass concentration values ($\mu\text{g}/\text{m}^3$) converted from particle counts using the manufacturer's default parameters. Temperature and humidity measurements (at sensor) are adjusted to reflect external temperature and relative humidity. | | | |

**(c) Health condition**
| **Instrument** | **Sampling frequency** | **Data format** | **Data types** |
| --- | --- | --- | --- |
| E-inhaler * | As needed | Excel | Inhaler usage records, including time, date, medication name (Ventolin), dose strength, event |
| E-spirometer ** | Daily | Excel | Spirometry test measures (with units), including date, time, FVC (L), FEV<sub>1</sub> (L), FEV<sub>6</sub> (L), FEV/FVC (%), FEF25-75 (L/s), and PEF (L/m) |
| Mi-Band | 15 minutes | CSV | Start date and time, end date and time, calories (kcal), distance (m), average/min/max speed (m/s), walking duration (s), running duration (s), step count, move minutes count |

**Notes:**
\* The e-inhaler records the following events, including “inhaler installed”, “inhaler removed”, and “medication”.
\*\* Each participant is required to perform three or more repeated tests during a short period in a day. Each spirometry test is marked with a quality assessment label, including “Good blow”, “Blow out faster”, “Blow out longer”, “Don’t hesitate”, “Abrupt end”, and “Don’t start too early”.

**(d) Health perception**
| Instrument | Sampling frequency | Data format | Data types |
| --- | --- | --- | --- |
| Paper-based survey | Weekly | CSV | Self-reported Asthma Control Test (ACT) |
| Paper-based survey | Weekly | CSV | Self-reported Asthma Quality of Life Questionnaire (AQLQ) |

### 2.2 Data Pre-processing

The PM exposure and meteorology data were sampled every minute by the portable sensor. In the raw tabular data, 19% of the rows consisted of at least one missing PM value (PM_1.0_, PM_2.5_, or PM_10_). We used an iterative imputer (Bayesian Ridge Regression) [6] to fill in the missing PM values based on the relationship between one feature and the others, utilizing the available data points, including PM_1.0_, PM_2.5_, PM_10_, temperature, and relative humidity. After missing data imputation, PM exposure and meteorology values were aggregated into hourly median values to reduce the impact of extreme values at the minute level. Hourly values were further aggregated into daily mean values to represent the daily average air pollution exposure.

Moreover, spirometry tests were performed by the study participants daily (at least three repeated tests during a short period). We only selected spirometry tests with “good blow” labels to reduce the potential measurement errors. Spirometry values were aggregated into daily maximum values.

Further, we calculated the mean and standard deviation of the daily PM and spirometry values. Any values out of the normal range (mean ± 3 × standard deviation) were considered outliers and removed.

### 2.3 Statistical Analysis

After data pre-processing, we performed a preliminary analysis to examine the relationship between personal air pollutant exposures and personal health conditions and health perceptions. The key variables are listed as follows.

Health conditions refer to objective assessments of one’s own health. For health conditions, the primary outcomes are the two lung function indicators that have been widely used in previous medical studies: FEV_1_, which measures how much air one can exhale during a forced breath during the first second, and FVC, which measures the total amount of air exhaled during the FEV test [7]. In addition, the secondary outcome is the number of asthmatic medications received by the subject as recorded by the e-inhaler.

Health perceptions refer to subjective assessments of one’s own health and well-being. For health perception, the primary outcomes are the ACT and AQLQ scores measured weekly. The ACT score is calculated using a standardized self-reported questionnaire that measures how well the symptoms of asthma are controlled. The AQLQ score is calculated using a standardized self-reported questionnaire that evaluates the quality of life of asthmatic patients.

For personal air pollutant exposures, PM_2.5_ and PM_10_ were selected due to their well-documented adverse health impacts in HK [8]. Moreover, PM_1.0_ was included in our analysis, given that the adverse health impacts of PM_1.0_ may be stronger than PM_2.5_ [9]. Further, demographic information, including age (number of years), gender (male or female), and body mass index (BMI), and meteorology information, including temperature (degree Celsius) and relative humidity (%), were included as covariates [10-12]. BMI (kg/m^2^) is calculated as the body weight divided by the square of the body height.

For each health condition and perception outcome, a univariate linear regression analysis was performed to examine the relationship between the personal air pollutant exposure variable and the outcome variable. A multivariate linear regression analysis was also performed to examine the statistical relationship after controlling for covariates. All variables were measured at the daily level (except for the three demographic variables). Single-pollutant models were used due to the collinearity between PM_1.0_, PM_2.5_, and PM_10_. Two-sided *p*-value < 0.05 was considered statistically significant.

## 3. Preliminary Results and Discussions

### 3.1 Preliminary Results

After data pre-processing, 44 daily observations were obtained. Table 2a shows the characteristics of the study subjects. Table 2b shows the descriptive statistics of the exposure and outcome variables after removing outliers, including the mean, standard deviation (SD), median, and interquartile range (IQR) values. The level of average PM exposure during the study period is relatively low, given that the study subjects often stayed at home (especially during the Covid-19 period) and indoor PM exposure is lower than outdoor levels in homes without strong PM sources.

**Table 2.** Descriptive statistics

(a) Characteristics of the asthma subjects
| Variable | Characteristic |
| --- | --- |
| Age: mean (range) | 13.1 (12-15) |
| Gender: percentage |  |
| Male | 50% |
| Female | 50% |
| BMI: mean (range) | 19.5 (16.3-24.4) |

(b) Descriptive statistics of daily exposure and outcome variables
| Variable (number of observations) | Mean ± SD | Median | IQR |
| --- | --- | --- | --- |
| 24-h average $PM_{1.0}$ ($n=43$) | $3.82 \pm 2.18$ | 3.38 | 1.98 |
| 24-h average $PM_{2.5}$ ($n=43$) | $4.48 \pm 2.39$ | 4.07 | 2.21 |
| 24-h average $PM_{10}$ ($n=44$) | $5.37 \pm 2.98$ | 4.64 | 3.38 |
| $FEV_1$ ($n=44$) | $2.61 \pm 0.61$ | 2.37 | 0.96 |
| FVC (*n*=44) | $3.23 \pm 0.81$ | 3.05 | 1.26 |

For the univariate relationship between personal air pollutant exposures and health conditions measured by FEV_1_ and FVC, we determined that all correlations are significant and negative, including PM_1.0_ exposure vs. FEV_1_ (R^2^=12%; *p*=0.023; Figure 1a), PM_1.0_ exposure vs. FVC (R^2^=15%; *p*=0.010; Figure 1b), PM_2.5_ exposure vs. FEV_1_ (R^2^=13%; *p*=0.019; Figure 1c), PM_2.5_ exposure vs. FVC (R^2^=16%; *p*=0.008; Figure 1d), PM_10_ exposure vs. FEV_1_ (R^2^=14%; *p*=0.012; Figure 1e), and PM_10_ exposure vs. FVC (R^2^=18%; *p*=0.005; Figure 1f).

**Figure 1.**
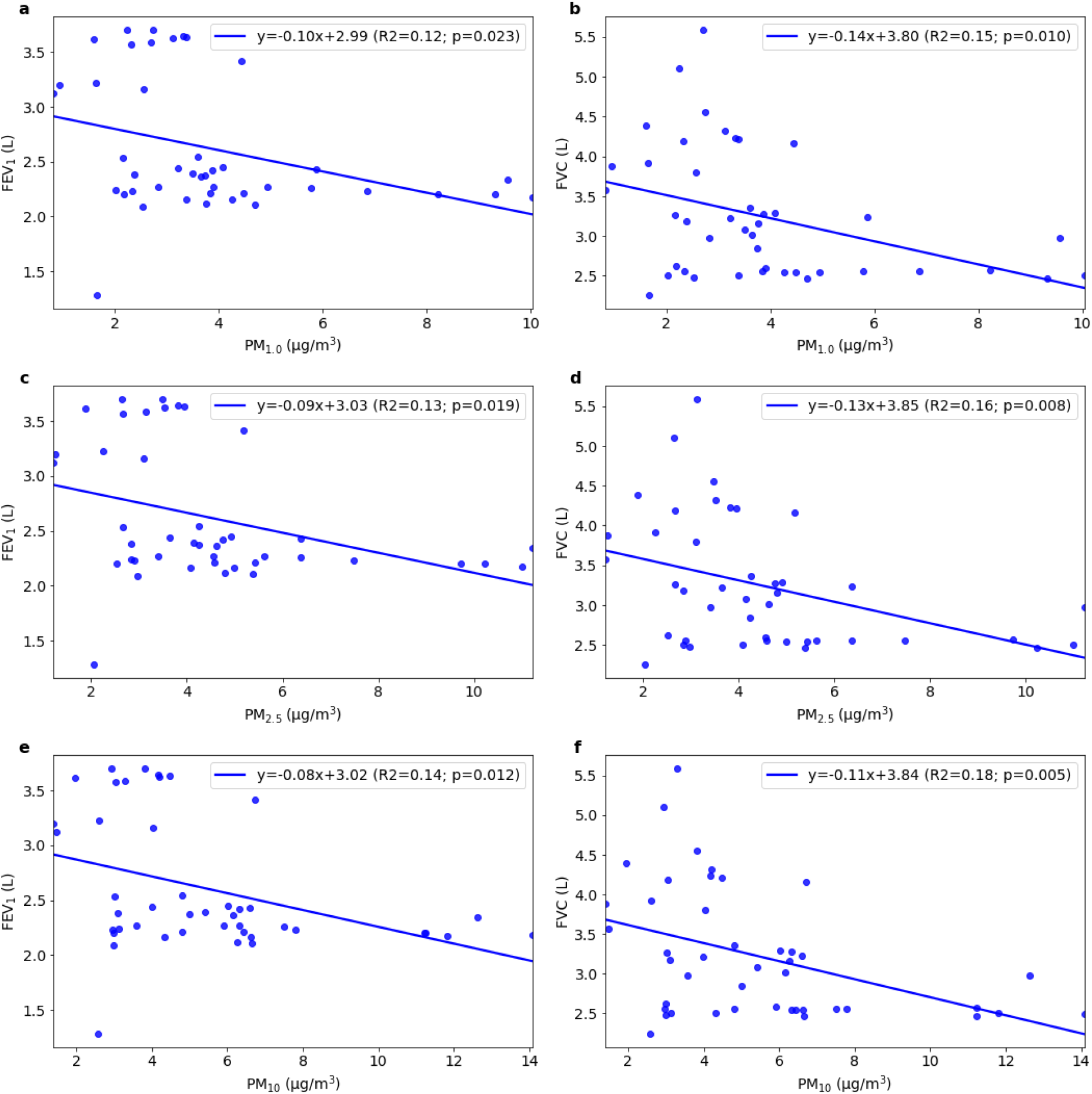
The univariate relationship between (a) daily personal PM_1.0_ exposure vs. FEV_1_, (b) daily personal PM_1.0_ exposure vs. FVC, (c) daily personal PM_2.5_ exposure vs. FEV_1_, (d) daily personal PM_2.5_ exposure vs. FVC, (e) daily personal PM_10_ exposure vs. FEV_1_, and (f) daily personal PM_10_ exposure vs. FVC

Moreover, after accounting for covariates, including age, gender, BMI, temperature, and relative humidity, we found a significant relationship between PM_1.0_ exposure vs. FVC (Coefficient=-0.1224; *p*=0.032; Table 3a), PM_2.5_ exposure vs. FVC (Coefficient=-0.1177; *p*=0.021; Table 3b), PM_10_ exposure vs. FEV_1_ (Coefficient=-0.0703; *p*=0.019; Table 3c), and PM_10_ exposure vs. FVC (Coefficient=-0.1204; *p*=0.006; Table 3d). However, the following two statistical correlations are negative and insignificant after controlling for covariates: PM_1.0_ exposure vs. FEV_1_ (Coefficient=-0.0679; *p*=0.084) and PM_2.5_ exposure vs. FEV_1_ (Coefficient=-0.0622; *p*=0.059).

**Table 3.** The significant relationship between PM exposure and spirometry after adjusting for covariates

**(a) PM<sub>1.0</sub> vs. FVC**
| **Dependent Variable: FVC** |  |  |  |
| --- | --- | --- | --- |
| **Number of Observations: 43** |  |  |  |
| **Adjusted R<sup>2</sup>: 38.6%** |  |  |  |
| **Variable** | **Coefficient** | **95% Confidence Interval** | ***p*-value** |
| Intercept | 5.1497 | [-2.283, 12.583] | 0.169 |
| Age | -0.3435 | [-1.183, 0.496] | 0.412 |
| Gender [=male] | -1.1810 | [-2.798, 0.436] | 0.147 |
| BMI | 0.4250 | [0.036, 0.814] | 0.033 * |
| Temperature | -0.1523 | [-0.368, 0.063] | 0.160 |
| Relative humidity | -0.0035 | [-0.019, 0.012] | 0.661 |
| PM<sub>1.0</sub> exposure | -0.1224 | [-0.234, -0.011] | 0.032 * |
| * $p$-value < 0.05 | | | |

**(b) PM<sub>2.5</sub> vs. FVC**
| **Dependent Variable: FVC** |  |  |  |
| --- | --- | --- | --- |
| **Number of Observations: 43** |  |  |  |
| **Adjusted R<sup>2</sup>: 39.8%** |  |  |  |
| **Variable** | **Coefficient** | **95% Confidence Interval** | ***p*-value** |
| Intercept | 5.2988 | [-2.026, 12.624] | 0.151 |
| Age | -0.3559 | [-1.187, 0.476] | 0.391 |
| Gender [=male] | -1.2325 | [-2.838, 0.373] | 0.128 |
| BMI | 0.4340 | [0.049, 0.819] | 0.028 * |
| Temperature | -0.1554 | [-0.366, 0.056] | 0.144 |
| Relative humidity | -0.0033 | [-0.019, 0.012] | 0.674 |
| PM<sub>2.5</sub> exposure | -0.1177 | [-0.217, -0.019] | 0.021 * |
| * $p$-value < 0.05 | | | |

**(c) PM<sub>10</sub> vs. FEV<sub>1</sub>**
| **Dependent Variable: FEV<sub>1</sub>** |  |  |  |
| --- | --- | --- | --- |
| **Number of Observations: 44** |  |  |  |
| **Adjusted R<sup>2</sup>: 51.1%** |  |  |  |
| **Variable** | **Coefficient** | **95% Confidence Interval** | **$p$-value** |
| Intercept | 1.9995 | [-2.986, 6.985] | 0.422 |
| Age | -0.0341 | [-0.596, 0.528] | 0.903 |
| Gender [=male] | -0.2939 | [-1.396, 0.809] | 0.592 |
| BMI | 0.2400 | [-0.023, 0.503] | 0.072 |
| Temperature | -0.1125 | [-0.253, 0.028] | 0.112 |
| Relative humidity | 0.0008 | [-0.010, 0.011] | 0.885 |
| PM<sub>10</sub> exposure | -0.0703 | [-0.128, -0.012] | 0.019 * |
| * $p$-value < 0.05 | | | |

**(d) PM<sub>10</sub> vs. FVC**
| **Dependent Variable: FVC** |  |  |  |
| --- | --- | --- | --- |
| **Number of Observations: 44** |  |  |  |
| **Adjusted R<sup>2</sup>: 43.9%** |  |  |  |
| **Variable** | **Coefficient** | **95% Confidence Interval** | **$p$-value** |
| Intercept | 6.1363 | [-0.985, 13.257] | 0.089 |
| Age | -0.4229 | [-1.226, 0.380] | 0.293 |
| Gender [=male] | -1.4506 | [-3.026, 0.124] | 0.070 |
| BMI | 0.4782 | [0.103, 0.853] | 0.014 * |
| Temperature | -0.1800 | [-0.380, 0.020] | 0.077 |
| Relative humidity | -0.0021 | [-0.017, 0.013] | 0.779 |
| PM<sub>10</sub> exposure | -0.1204 | [-0.204, -0.037] | 0.006 * |
| * $p$-value < 0.05 | | | |

The statistical relationship between personal air pollutant exposure and asthma medication is yet to be conclusive due to the lack of data. Observations on the total number of e-inhaler doses are very limited. Similarly, the number of health perception data points is too limited. With regard to the asthma control survey, very little about the symptoms of the asthmatics has been reported over the study period. With regard to the asthma-related quality-of-life survey, the quality-of-life indicators show very little variation. In view of this, we have put forward new practical suggestions on how to motivate our young asthmatic subjects to respond more actively to the health condition and perception surveys.

### 3.2 Discussions

Our preliminary results show that a higher level of PM_1.0_, PM_2.5_ and PM_10_ would deteriorate the health conditions of young asthmatics in HK, as shown by a lower FEV_1_ and a lower FVC value. This is the first study to investigate the relationship between PM_1.0_ and FEV_1_ and FVC of young asthmatics in Hong Kong, based on personal exposures obtained from portable sensors.

Results of our pilot study also provide insights on the proper effect size of our follow-up study. Following the previous studies on personal air pollutant exposure and lung function [10-12], we have adjusted the regression coefficients based on one IQR change in PM_2.5_ exposure level. For one IQR change in personal PM_2.5_ exposure, the change in FEV_1_ and FVC was -0.137 and -0.260, respectively. We found that the effect size in our study, as represented by the adjusted regression coefficient, is larger than the previous studies (Table 4).

**Table 4.** A comparison of the effects of personal air pollutant exposures on lung functions

| Subject Selection and Sample Size | Outcome | Personal Exposure | Coefficient * | Reference |
| --- | --- | --- | --- | --- |
| 36 healthy college students | FEV<sub>1</sub> | PM<sub>2.5</sub> | -0.011 | [10] |
| 36 healthy college students | FVC | PM<sub>2.5</sub> | -0.007 | [10] |
| 53 asthmatic schoolchildren (aged 9 to 18 years old) | FEV<sub>1</sub> | PM<sub>2.5</sub> | -0.059 | [11] |
| 43 asthmatic children (aged 5 to 13 years old) | FEV<sub>1</sub> | PM<sub>2.5</sub> | -0.024 | [12] |
| 43 asthmatic children (aged 5 to 13 years old) | FVC | PM<sub>2.5</sub> | -0.017 | [12] |
| 9 asthmatic schoolchildren (aged 12 to 15 years old) | FEV<sub>1</sub> | PM<sub>2.5</sub> | **-0.137** | Pilot study |
| 9 asthmatic schoolchildren (aged 12 to 15 years old) | FVC | PM<sub>2.5</sub> | **-0.260** | Pilot study |
| **Notes:**<br>* To compare the effects of personal air pollution on health outcomes across different studies, the regression coefficient was calculated as the change in outcome per one IQR change in air pollution level after adjusting for covariates [10-12]. |  |  |  |  |

Further, using the pilot study data, we have performed a power analysis to estimate the sample size for our follow-up main study. The primary null hypothesis is that personal PM exposure would not change the FEV_1_ and FVC of young asthmatics in HK. To better measure the local effect size of personal air pollution exposure in a multivariate regression context, we calculated Cohen’s *f*^*2*^ value [13] based on (1) the adjusted R^2^ value of each regression model shown in Table 3 and (2) the adjusted R^2^ value of the corresponding regression model without the PM exposure variable. We found that the Cohen’s *f*^*2*^ value ranges from 0.107 to 0.200. Then, using the effect size of 0.107, we performed a two-sided F test under a multivariate linear regression setting (three predictors to be tested and ten predictors in total) [14] to calculate the sample size that gives 80% power at a 5% significance level. We found that the lowest number of asthmatic subjects is 107. After accounting for a potential non-compliance and dropout rate of 10% to 15% (given that children in HK normally show much higher compliance), we determined that 125 asthmatic subjects are sufficient for our main study. Moreover, to obtain more rigorous statistical findings by comparing the difference between the asthmatic subjects and healthy subjects, 125 healthy subjects will also be enrolled in our main study in a 1:1 ratio.

Our follow-up main study will collect more longitudinal health condition and perception and air pollutant exposure data from 125 asthmatic subjects and 125 healthy subjects. This longitudinal dataset will help us better investigate the statistical relationship between personal air pollutant exposure and health condition and perception, by utilizing more advanced statistical modelling to account for (1) the repeated measures of health outcomes and (2) the lag effects of personal air pollutant exposure. We expect to obtain more rigorous statistical findings in our upcoming main clinical study.

## 4. Major Challenges of the Pilot Study

As hospitals and clinics have focused on combatting COVID-19, the recruitment of subjects has been adversely affected. The Hospital Authority has required all non-urgent cases to stay away from hospitals to minimize risks. Additional challenges in data collection and statistical analysis are listed below (See solutions to overcome such challenges in Section 5).

### 4.1 Missing Data

Notable missing values have been observed in the data collected from different types of sensors. During the pilot study, the missing data can be due to human errors (e.g., the subject may forget to perform the required actions) or data transmission errors (e.g., the sensor/phone may fail to connect to the internet).

### 4.2 Automated Data Collection

The data collection process is yet to be fully automated and monitored. In the pilot study, the data collected from the e-inhaler, e-spirometer, and Mi-Band were downloaded and processed manually before further analysis. Moreover, the questionnaires are yet to be automated and conducted online.

### 4.3 Statistical Modeling for Longitudinal Data

Due to the small number of data points, advanced statistical models for longitudinal data, such as linear mixed-effect models, are yet to be utilized.

## 5. Follow-up Actions and Directions

The major challenges of the pilot study are interlinked. By improving our data collection procedure and engaging with more participants, we expect to establish a high-quality longitudinal dataset for advanced statistical analysis. More specifically, to overcome these challenges, our follow-up work can be divided into two main directions: (1) improving data collection and analysis and (2) increasing and mobilizing subject engagement.

### 5.1 Data Collection and Analysis

#### 5.1.1 Building an Integrated Data Management Platform

We are building an integrated data management platform to automatically collect, process, monitor, and manage the data across all sources, utilizing a cloud computing server equipped with strong measures for user privacy and data security. So far, the portable sensor data collection procedure has been successfully integrated into the data management platform. Moreover, an alert system, as a part of the data management platform, will be deployed to identify and fix data synchronization and transmission issues, and issue missing data alerts to (1) inform the study team to take action and (2) remind the subjects to follow the data collection procedure closely.

#### 5.1.2 Calibrating Air Pollution Data

Calibrating air pollution data can reduce the measurement errors and allow the subjects to better understand the risks of his/her exposure. In the portable sensor, PM pollutants are measured by a laser particle counter. The PM sensor can be calibrated based on the raw data (bin counts for a range of particle size bins) while accounting for meteorology factors [15]. Our sensor calibration can be performed seasonally to capture the seasonal variation of air pollution in HK (for more details, please refer to our sensor calibration report).

#### 5.1.3 Integrating E-surveys into the UMeAir App

We are integrating the e-surveys into the UMeAir App and the data management platform, based on a separate online e-survey system developed by us previously. The UMeAir App will motivate the subjects to fill in the surveys on time with gentle musical alerts. We will also look for more incentive and empowerment measures to our young participants, along the line of good citizenship recognition/differentiation; such as running smart competitions for our participants with chances of visiting our new HKU innovation facilities online, opportunities of visiting online new high-tech start-ups in HK or Israel, inviting our participants to our UMeAir Club as VIPs, and awarding certificates of green, healthy and smart citizenship to our VIPs, in order to improve the quality of questionnaire data collection.

#### 5.1.4 Improving the Statistical Analysis

After collecting more longitudinal data with higher data integrity and fewer missing values, we aim to improve the current statistical methodology. We will adopt advanced statistical models (such as the linear mixed-effect models) to account for the repeated measurements and the lagged effects of air pollutant exposure, while controlling for important confounders and autocorrelation in longitudinal data. We will also address non-linearity and multicollinearity if any are found in our follow-up study. We expect to obtain more rigorous statistical findings in our follow-up study.

### 5.2 Subject Engagement and Recruitment

#### 5.2.1 Troubleshooting Equipment Usage in Close Collaboration with QMH

During the pilot study, considerable data gaps for most of the sensors and measurements were identified. While data collection automation will help, increasing the accessibility of the sensors and reducing technical barriers of the subjects can improve the engagement of the subjects and thus the integrity of the data. We are working closely with QMH staff to produce a series of instructional videos for the usage of different sensors for the study and provide timely technical assistance to both the staff and subjects.

#### 5.2.2 Recruitment of Healthy Subjects

In order to provide an effective control group, we are currently proceeding with the recruitment of healthy subjects for the pilot study. However, due to the suspension of schools as a result of the ongoing COVID-19 pandemic, there are difficulties in the recruitment of students. We are working closely with schools and starting to install air pollution sensors in schools. Meanwhile we shall recruit the healthy subjects from schools.

## 6. Conclusion

This report provides a summary of the current progress made in the TBR Air Pollution Clinical and Quality of Life Study. We determine that all correlations are significant and negative, including PM_1.0_ exposure vs. FEV_1_ (R^2^=12%; *p*=0.023; Figure 1a), PM_1.0_ exposure vs. FVC (R^2^=15%; *p*=0.010; Figure 1b), PM_2.5_ exposure vs. FEV_1_ (R^2^=13%; *p*=0.019; Figure 1c), PM_2.5_ exposure vs. FVC (R^2^=16%; *p*=0.008; Figure 1d), PM_10_ exposure vs. FEV_1_ (R^2^=14%; *p*=0.012; Figure 1e), and PM_10_ exposure vs. FVC (R^2^=18%; *p*=0.005; Figure 1f). Moreover, after accounting for covariates, including age, gender, BMI, temperature, and relative humidity, we found a significant relationship between PM_1.0_ exposure vs. FVC (Coefficient=-0.1224; *p*=0.032; Table 3a), PM_2.5_ exposure vs. FVC (Coefficient=-0.1177; *p*=0.021; Table 3b), PM_10_ exposure vs. FEV_1_ (Coefficient=-0.0703; *p*=0.019; Table 3c), and PM10 exposure vs. FVC (Coefficient=-0.1204; *p*=0.006; Table 3d). Further, using the pilot study data, we have performed a power analysis to estimate the sample size for our follow-up main study. Based on the primary null hypothesis is that personal PM exposure would not change the FEV_1_ and FVC of young asthmatics in HK, the lowest sample size that gives 80% power at a 5% significance level is 107. Hence, the sample size (or the total number of participated asthma subjects) expected for the follow-up longitudinal clinical study should be 125 (after adjusting for the non-compliance and withdrawal of subjects). Our pilot study has demonstrated the feasibility of research into the effects of personal air pollutant exposure on health condition and health perception. Our follow-up study will address the challenges identified in the pilot study, based on the proposed follow-up actions for subject engagement, data collection, and data analysis.

## Data Availability

Figure 1 and Table 1-4 have all associated raw data available on the group's Microsoft Azure server that will be released on a later date.

## Acknowledgement

We acknowledge the assistance of Miss Jennifer Lam, Research Nurse, Department of Pediatrics, Queen Mary Hospital, Hong Kong, in training recruited asthma subjects in using the medical and air pollution sensor devices, and in collecting clinical trial data. We also thank Mr. Andong Wang for his research assistance in this clinical trial project.

## Notes

### Competing Interest Statement

The authors have declared no competing interest.

### Funding Statement

This work is supported by the Theme-based Research Scheme of the Research Grants Council of the Hong Kong SAR Government, under Grant No. T41-709/17-N.

### Author Declarations

Institutional Review Board of The University of Hong Kong/ Hospital Authority Hong Kong West Cluster.

